# Early epidemiological and clinical manifestations of COVID-19 in Japan

**DOI:** 10.1101/2020.04.17.20070276

**Authors:** Muhammad Qasim, Muhammad Yasir, Waqas Ahmad, Minami Yoshida, Muhammad Azhar, Mohammad Azam Ali, Chris Wang, Maree Gould

## Abstract

**Background:** Severe acute respiratory syndrome coronaviruses -2 (SARS-COV2) named as COVID-19 had spread worldwide and leading to 1,210,956 confirmed cases and 67,594 deaths

**Methods:** A data of 1192 confirmed cases and 43 deaths due to COVID-19 in Japan collected from the Ministry of Health, Labour and Welfare of Japan and analysed for different epidemiological parameters and their clinical manifestations. We used Clauset-Newman-Moore (CNM) clustering algorithm to develop web-network of confirmed cases to identified clusters of community transmission.

**Results:** Out of 1192 confirmed cases, 90.60% were symptomatic and 9.39% were asymptomatic. The prevalence of COVID19 in males was 56.29% and 43.20 % in females. The mean interval (±SD) from symptom onset to diagnosis was 6±22.6 days while mean interval (±SD) from contact to onset of symptoms was 5±19.5 days. People of age range 40-79 were more infected and deaths median age was 80. The main symptoms were fever, dry cough, fatigue and pneumonia. The main infected cities were Tokyo (195/1192, 16.35%), Hokkaido (160/1192 13.42%), Aichi (150/1192, 12.58%) and Osaka (145/1192, 12.16%). Only 2.34% cases had travel history from Wuhan China and Osaka music concert was identify as main cluster for community transmission. While 556 (46.64%) cases were clinically diagnosed and 557 (46.72%) were confirmed by using RT-PCR.

**Conclusions:** Other than, declare emergency Japan need to change their approach of diagnosing COVID-19, as asymptomatic cases prevalence is high and maybe it is reason for current sudden increase of cases. Screening centre should be establish away from hospitals, which are treating positive cases.

## Introduction

As the COVID-19 pandemic insidiously infiltrates and the numbers of infected people and deaths skyrocket in various locations around the world, an ongoing puzzle has been the comparatively slow rise of those numbers in Japan.^1^ But since last week, the numbers in Japan have begun to climb in a concerning way.^2^ Until April 7, 2020, there were 3,654 confirmed cases with 73 deaths reported. These cases exist other than the 712 laboratory-confirmed cases on the Diamond Princess cruise ship.^2,3^ On the other hand according to WHO the worldwide cases of COVID-19 reached to 1,051,635 with 56,985 deaths.^2^ 1^st^ case of COVID-19 reported in Japan on 15^th^ January, 2020 and he was returned from Wuhan, China the mainland from where disease started.^4^ On February 1, a passenger who disembarked from the Diamond Princess days earlier in Hong Kong tested positive for the COVID-19 coronavirus.^3^ The ship was quarantined immediately after it arrived in Japanese waters on February 3 with 3,711 passengers and crew members on board.^5^ Over the next month, more than 700 people on board became infected including a nurse and for several weeks, the ship remained the site of the largest outbreak outside of China.^6^ The outbreak of this disease has caused the Japanese government to take drastic measures to contain the outbreak, including the quarantine of millions of residents in Hokkaido, the Diamond Princess cruise ship and other affected cities. By March 24, 2020, there were 1192 confirmed cases of COVID-19 in Japan. In this study, we analysed the early epidemiological dynamics and clinical manifestations of these confirmed 1192 cases to understand the characteristics of COVID19 pandemic in Japan.^4^

The epidemiological analysis of the COVID19 pandemic provides a unique opportunity to characterize its transmission, source of community spread and the effectiveness of screening processes.^7^ While clinical manifestation of COVID19 cases will help clinicians to understand the dynamics of COVID-19 and help them to diagnose asymptomatic cases. The careful monitoring of cases and demographic data from the general community enables inferences that are critical to modelling the course of the outbreak, that are difficult to make during a widely disseminated epidemic.^8,9^ We highlighted the critical transmission parameters, such as; contact of people with an infected person, places that they visited before infected, demographic data and prevalence of symptoms. This study will help future monitoring of COVID-19 in particularly in Japan and generally worldwide to guide respective measures to contain its spread.

## Methods

Data of total 1192 confirmed and 43 deaths has been collected from 15^th^ January to the 24^th^ March 2020 from daily reports published by the Ministry of Health, Labour and Welfare of Japan.^4^ We analysed the data for demographic distribution of cases and epidemiological parameters such as clusters based upon hospitalisation, travel and contact with positive case history of confirmed cases. We also analysed the clinical manifestation of confirmed cases such symptoms, mean intervals time from onset of symptoms until confirmation of COVID19 presence and mean intervals from patients first contact with positive case to onset of symptoms. In order to understand the community transmission pattern and identify the hotspot for spread of COVID-19 cases in Japan. We ran a Clauset-Newman-Moore (CNM) clustering algorithm^10^. The Clauset-Newman-Moore algorithm forms a cluster on the basis of modularity, that is a degree of graphing network which divides the network into groups in such a way that there are many edges within the groups but few between the groups, thus identifying modular clusters. This clustering helped us to find the web of community transmission and highlight the main nodes points. We also analysed the hospitalisation history of confirmed cases to find any possible link of community spread of COVID-19 with health centres. As Japan, did not observed the complete lock down so we analysed the reason for rapid increase of cases in Japan in first week of April 2020 by looking into contact history of new cases with previously positive or asymptomatic cases.^11^ We also analysed the number confirmed cases, clinically diagnosed, laboratory confirmed or diagnosed based on CT scan and X ray.

## Results

The data analysis of total 1192 cases showed that 1080 (90.60%) cases were symptomatic and 112 (9.39%) were asymptomatic as shown in **Table 1**. Out of 1192 confirmed cases, 671 (56.29%) were male and 515 (43.20%) were female and rest status was unknown. While out 112 asymptomatic cases 105 (93.75%) were within country while 7 (6.25%) were in government charted flight came from Japan. While only 28 (2.34%) cases had travel history from Wuhan, China and only 21 (1.80%) cases were linked with Diamond Princess Cruise ship.^12^ The mean interval (±SD) from symptom onset to diagnosis was 6±22.6 days while mean interval (±SD) from first contact with positive case to onset of symptoms was 5±19.5 days. (**Table 2**) It has found that the highest number of patients infected with COVID-19 in Japan have ages ranging between 40-79, as shown in **Figure 1**. While 69.76% (30/43) of death were male and majority of deaths age range (70-90) (**Figure 1**). Young people with ages ranging from 20-40 are the least effected. In plotting the symptoms of COVID19 symptomatic (1080) infected patients, the results showed that fever (857/1080, 79.35%), cough (459/1080, 42.50%) and fatigue (316/1080, 29.25%) were the main symptoms found in patients, as shown in **Figure. 2**. While only 240/1080 (22.22%) patients showed pneumonia symptoms while 223/1080 (20.64%) showed respiratory or shortness of breath, as shown in **Figure.2**.

**Table 1:** Epidemiological parameters of COVID-19 confirmed cases until March 24, 2020 in Japan

| **Parameter** | **Cases**<br>**Number** | **Within**<br>**Country**<br>**(Japan)** | **Govt.**<br>**Chartered**<br>**flight**<br>**from**<br>**China** | **Data Not**<br>**Available** | **No**<br>**Travel**<br>**History**<br>**to**<br>**Wuhan** | **Linked with**<br>**Diamond**<br>**Princess Cruise**<br>**ship** | **Total** |
| --- | --- | --- | --- | --- | --- | --- | --- |
| **Symptomatic** | 1080<br>(90.60%) | - | - | - | - | - | 1080 |
| **Asymptomatic** | 112<br>(9.39%) | 105 | 7 | - | - | - | 112 |
| **Travel History**<br>**Wuhan(China)** | 28<br>(2.34%) | - | - | 9 | 1155 |  | 28 |
| **Were in**<br>**Diamond**<br>**Princess Cruise**<br>**ship** | 21<br>(1.76%) |  |  | 9 | 1162 |  | 21 |

**Table 2:** Clinical manifestation of COVID-19 confirmed cases until March 24, 2020 in Japan

Tables 2: Clinical manifestation of COVID-19 confirmed cases until March 24, 2020 in Japan
| **Parameter** | **Data**<br>**Available**<br>**(Complete**<br>**Data)** | **Data Not**<br>**Available** | **Mean (Days)** | **Total** |
| --- | --- | --- | --- | --- |
| **Meantime from**<br>**Onset of**<br>**Symptoms to**<br>**Confirmation** | 1000 | 192 | 6±22.6 | 1192 |
| **Meantime from**<br>**Contact with**<br>**COVID19**<br>**positive case to**<br>**Onset of**<br>**Symptoms** | 148 (112) | 1044 | 5±19.55 |  |

**Figure 1:**
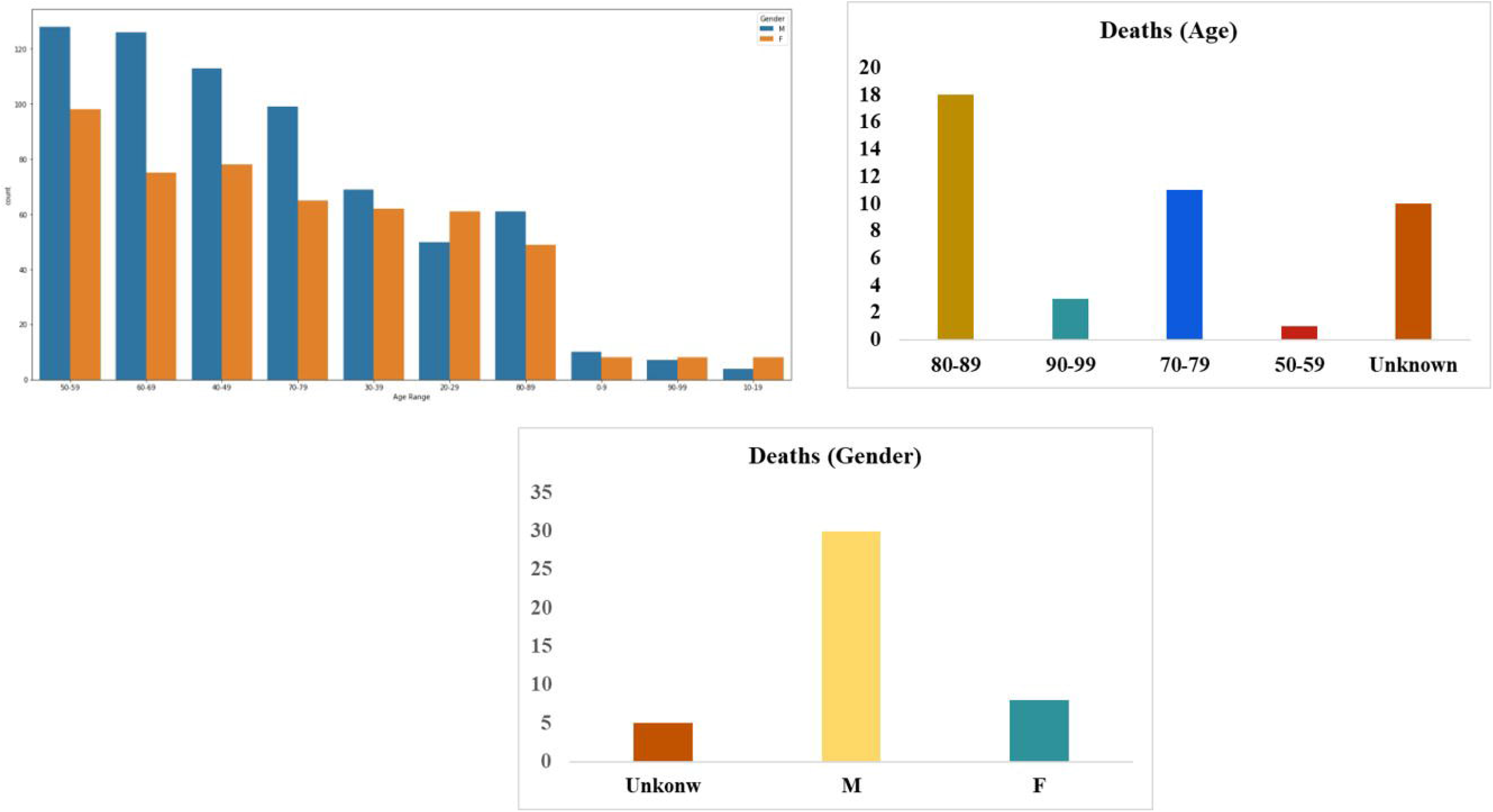
(a) Gender and age distribution of COVID-19 confirmed cases in Japan (b) Age distribution of deaths due to COVID-19 (c) Gender distribution of deaths due to COVID-19 in Japan until 24 March 2020

**Figure 2:**
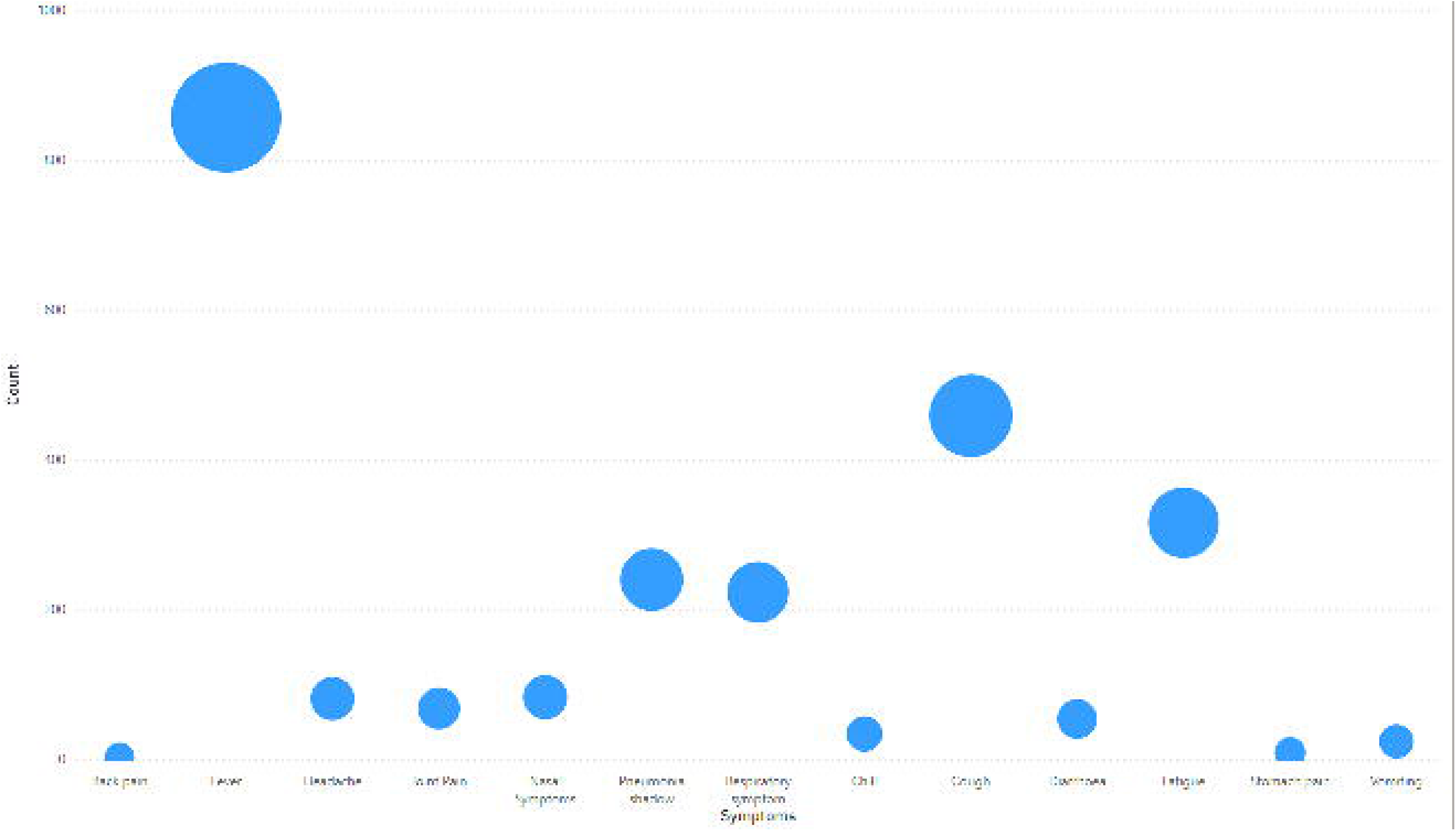
Graph of major symptoms appeared in symptomatic case of COVID-19 in Japan

The prevalence of COVID-19 in Japan until March 24, 2020 showed that; Tokyo (195/1192, 16.35%), Hokkaido (160/1192 13.42%), Aichi (150/1192, 12.58%), Osaka (145/1192, 12.16%) and Hyogo (114/1192, 9.56%) were top five most affected areas, as shown on map **Figure 3**.

**Figure 3:**
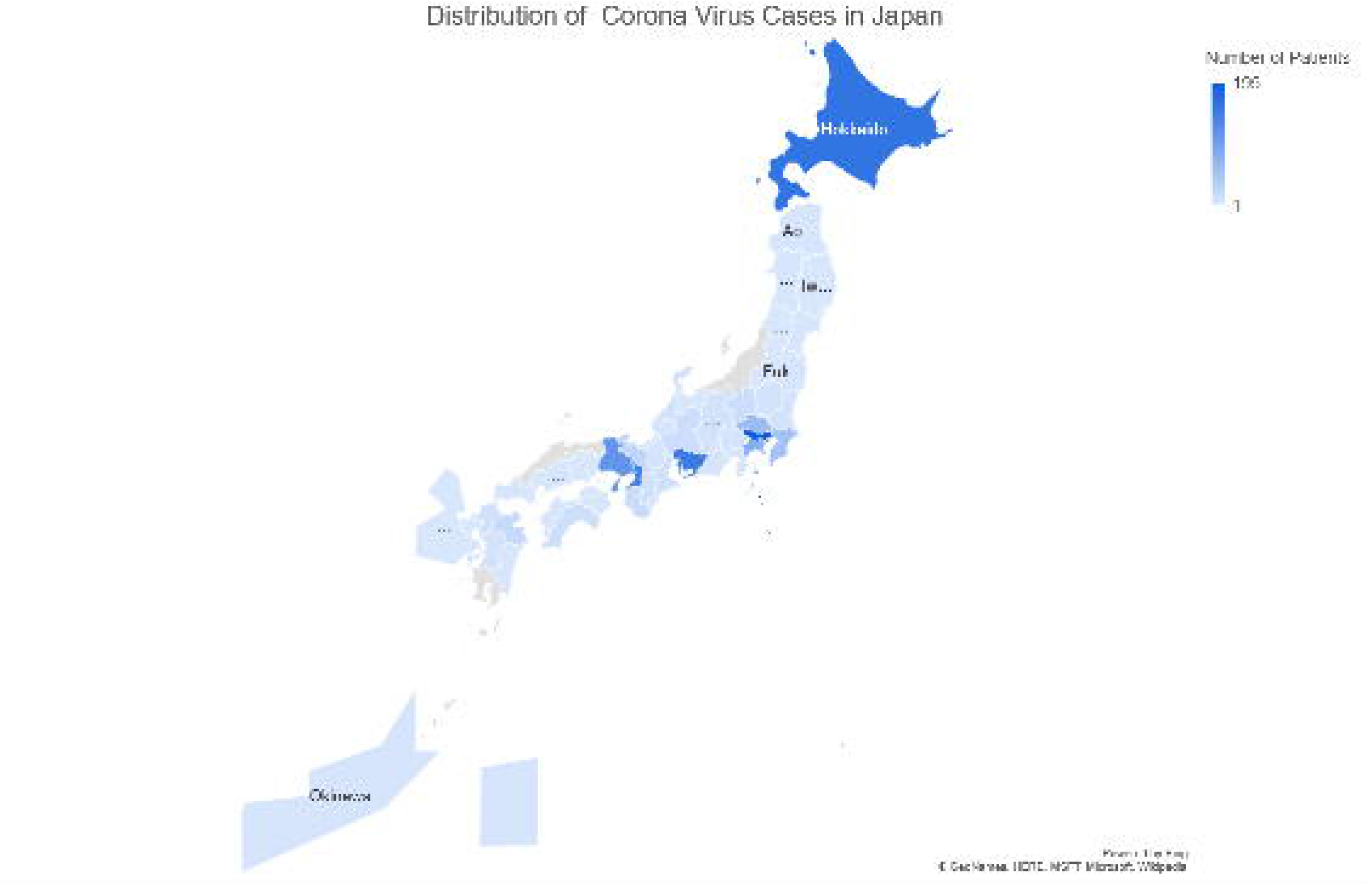
geographical distribution case of COVID-19 confirmed cases in Japan until March 24, 2020

The information about infected people’s occupations was available for only 87/1192 (7.26%) cases and it was discovered that 29/87 (33.33%) were doctors, paramedical staff and hospital workers as shown in **Figure 4**. An interesting fact observed in the case of Hokkaido City Hospital was about 50% of the casualties (i.e. a total of 3) occurred in the same hospital. Similarly, 50% of nurses (i.e. 3) were also treated in the same hospital and this hospital treated the highest number of infectious patients in Japan. So most probably, this hospital was also a source of transmitting the infection, as maybe those people who visited this hospital also became infected. Overall, the history of the patients showed that 90% of the infected patients had a previous hospitalisation history, as shown in **Figure 4**. Similarly, 60% of cases with a previous hospitalisation history had contact with COVID-19 positive cases. While monthly data showed that patients with a hospital visit history were fewer in start but then increased gradually, similarly COVID-19 confirmed cases with a history of patient contact showed a positive trend that increased each day that passed, which confirmed the community transmission of the virus.

**Figure 4:**
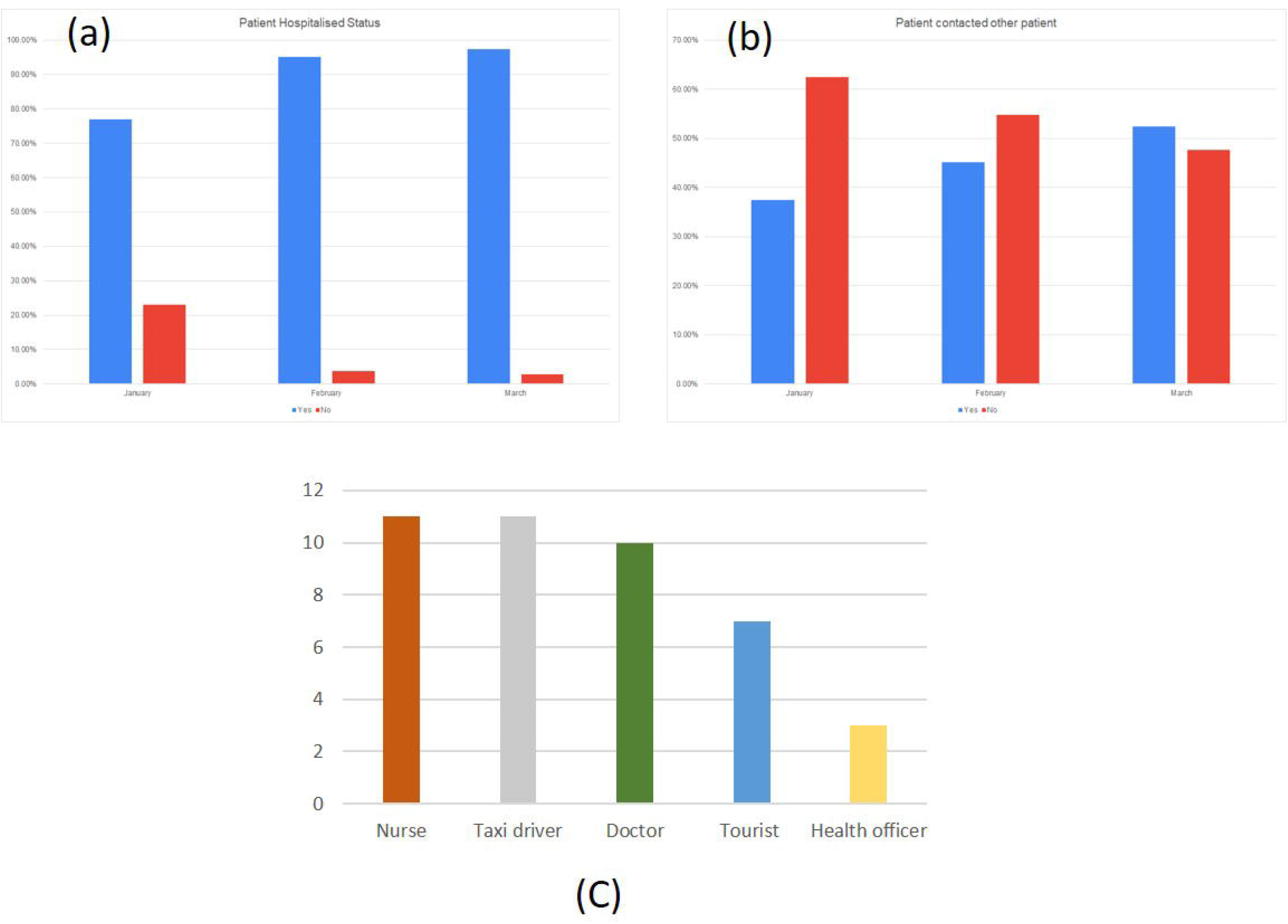
History of confirmed cases of COVID 19 (a) Pervious Hospitalization (b) has contact with positive case and (C) confirmed cases’s occupations distribution

We used the Clauset-Newman-Moore algorithm to find clusters and interlink the patients to find the centres of the community transmission (**Figures 5 and 6**). We have found that a patient A-34 from Aichi (age 60-69) with a travel history from Hawaii (USA), and patient A-347 from Aichi (age 80-89) had contact history with patient A-229 from the same city and patient A-391 from Hyogo greatly increased the spread of infection within the community. It was also found that a live music concert was the main point of community spread of COVID-19 in Osaka, as shown in **Figure 6**. Many people think that spread of infection was due to a Chinese tourist that came onto the cruise ship but our findings show that this was not the case.^12^

**Figure 5:**
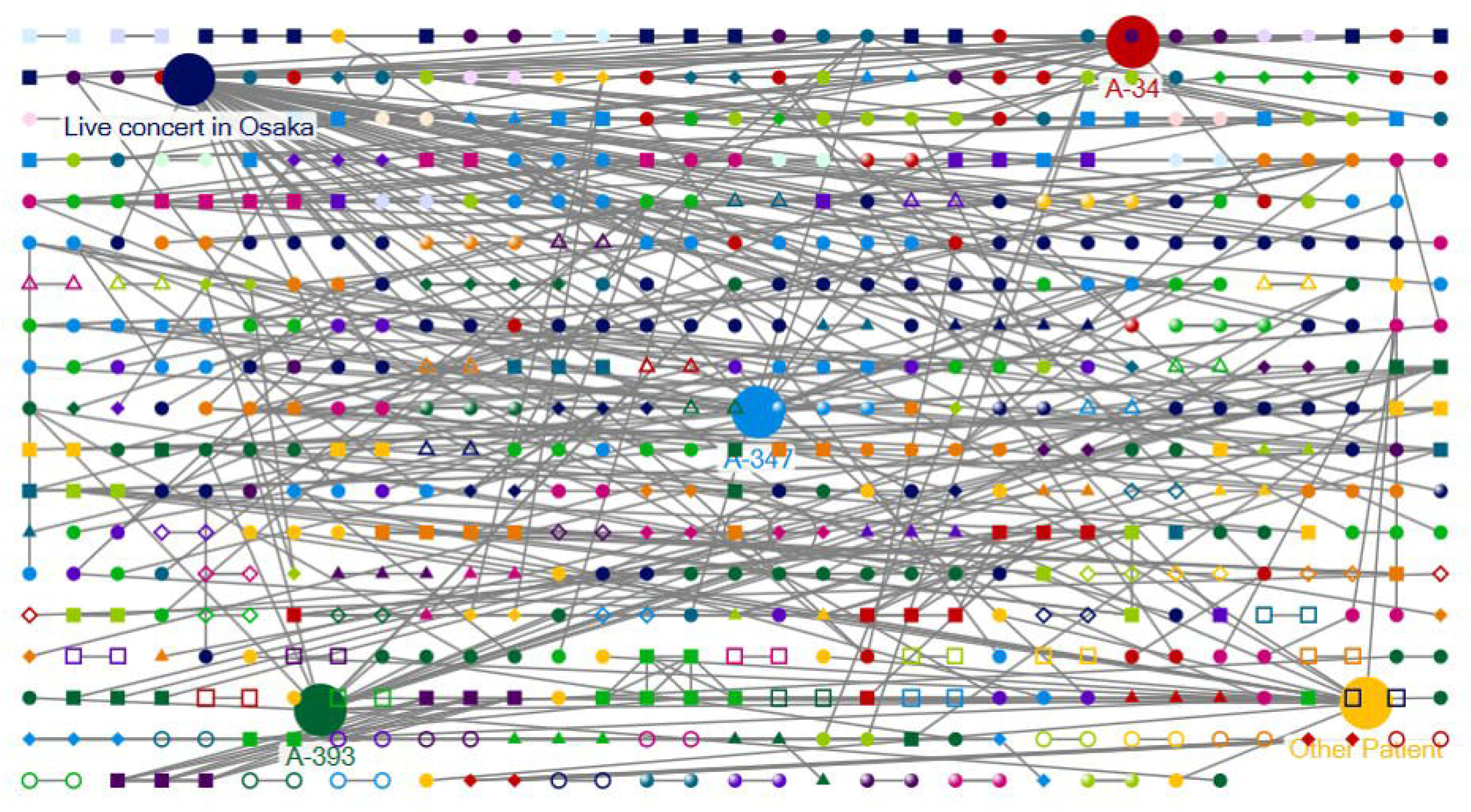
Clauset-Newman-Moore (CNM) clustering algorithm web-chart showing linkages of confirmed cases contact with pervious positive case of covid19 in Japan

**Figure 6:**
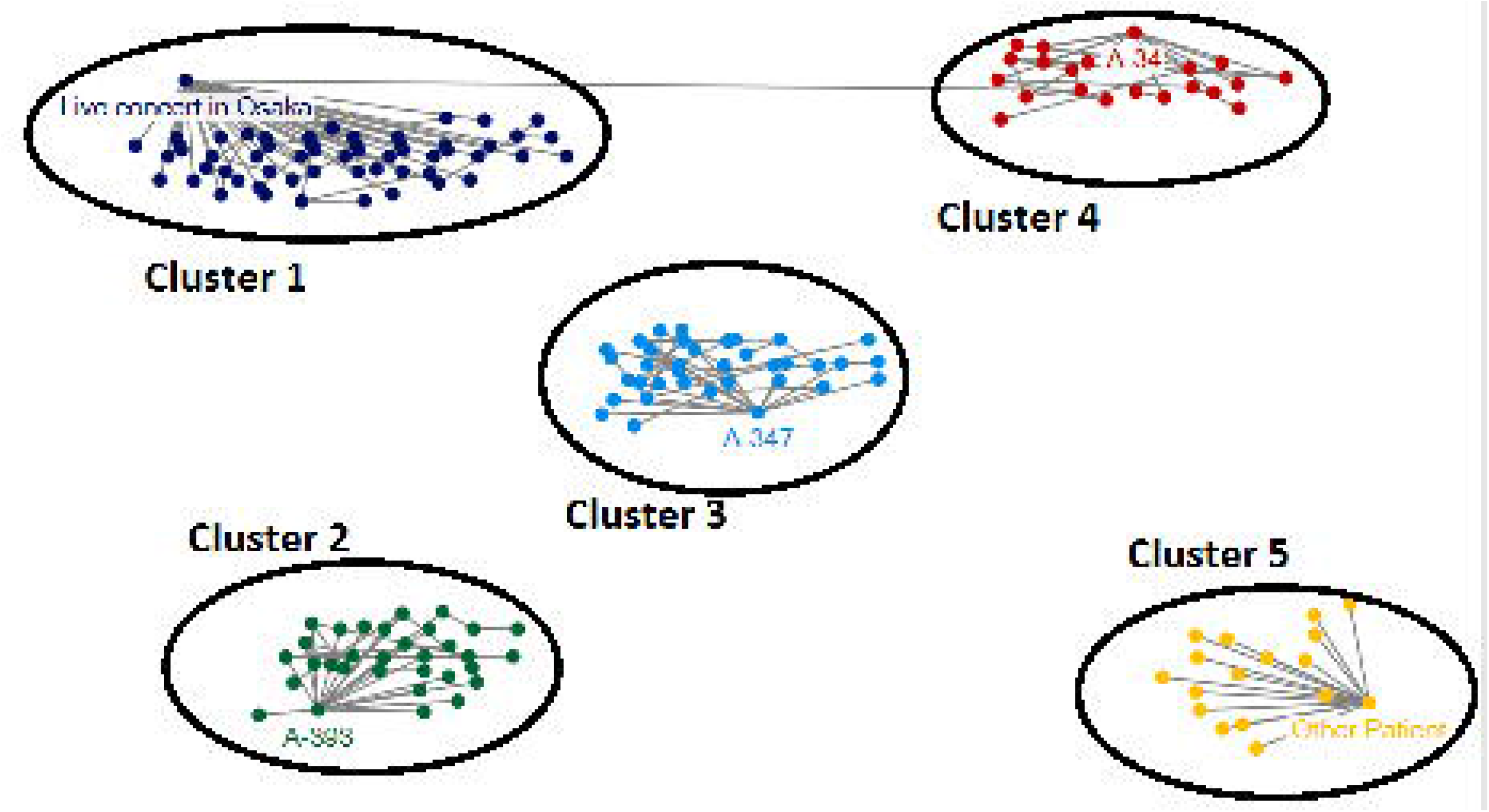
Clauset-Newman-Moore (CNM) clustering algorithm web-chart showing five main cluster of community transmission of COVID19 in Japan till 24^th^ March, 2020

The screening methodology of Japan indicated that they have a lower number of tests to confirm their patients diagnoses and instead rely upon a clinical diagnosis. As data showed that out of the 1192 patients, 556 (46.64%) cases were clinically diagnosed while almost same number of patients were confirmed using RT-PCR testing (46.72%) **Figure 7**. While the remaining cases were confirmed based upon X-ray or CT Scan. As clinically, it is hard for doctors to diagnose COVID-19 in asymptomatic patients, therefore there are chances many positive COVID-19 cases lost.

**Figure 7:**
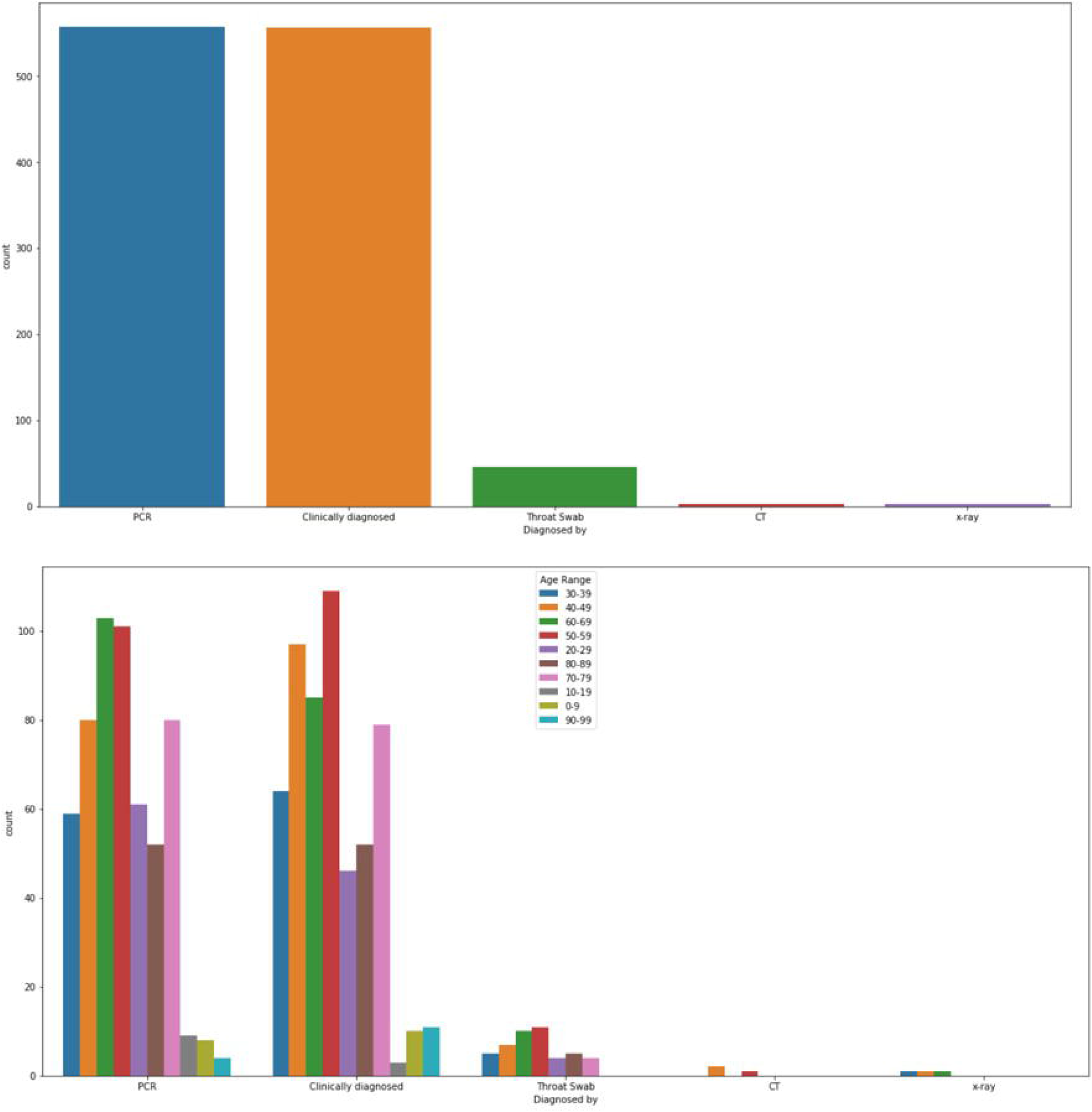
Confirmation of COVID-19 positive cases by different diagnostics methods

## Discussion

Our study results showed that the main characteristic of COVID-19 in positive patients in Japan were fever, fatigue and cough. While the main burden of COVID-19 cases were prevalent in the cities of Tokyo, Hakkadio, Aichi and OSKA. While data analysis showed that, the hospital was the prime suspect for community transmission of COVID 19. While the Clauset-Newman-Moore algorithm analysis showed that most of the reported cases have contact with positive cases within the country and a music concert in Osaka has played a major role for the community spread of COVID 19. Japan has an excellent public health care system.^13^ Care is affordable, so most people see a physician when they are beginning to feel ill, rather than when the conditions have worsened. So far, the low number of cases until the end of March can be directly related to the low number of test performed by Japan as they rely more on clinical diagnosis, X-ray and CT scan reports.^14^ As our this study showed that mean interval time for onset of symptoms is 5 days and in other reported studies it is 14-21 days for COVID-19 so there are many chances that many asymptomatic cases were missed and lost in community.^15^ While mean interval time from onset of symptoms to diagnostics in case of Japan was found 6±22.6 days while in case of Korea it was 4.3±4.1 days.^16^ The current sudden rise in COVID 19 cases in Japan now can be link to this fact.

With relation to CT scanners, Japan may have the best diagnostic imaging devices in the world, as the number per 100,000 people is 101 and their rely more on these second line diagnostics can pose a threat to their health system or methodology to handle COVID-19 cases.^17^ Therefore, one key reason for the low number of infections recorded, is that Japan has imposed strict criteria to be eligible for testing, focusing on giving tests only to people who have sustained fevers for more than four days, combined with overseas travel, close contact with an infected person or lung symptoms severe enough to warrant hospitalization. The goal of this approach has not been to identify all the infected people, but rather to focus resources on those most in need of treatment but it seems recent sudden rise in cases showed, this approach is not working effectively.

While our CNM) clustering algorithm web graph (Figure 5 and 6) showed that, lack of strict measure to hold big community gathering dent the Japan measures to mitigate the COVID-19 spread, as most of community transmission cases linked with music concert in Oska. Furthermore our analysis showed that there almost 10% cases were asymptomatic (Table 1) so if we model this number its projection shows an alarming picture for Japan.^18^ Now, Japan has imposed the state emergency imposed in the capital, Tokyo, and six other prefectures accounting for about 44% of Japan’s population from April 7 2020 until 6 May 2020 to press the public to stay home as compared to previous approach to trust on public obedience.^19^ But our study suggest they also need to revise their approach for diagnosis of COVID-19 and increase their laboratory based testing for screening of people at large scale. Second, they need to protect their health care workers and keep public away from hospital until they really need it. For screening of public sample collections centre must be set away from hospitals where positive cases are under treatments.

## Conclusion

Out study showed that people of age above 40 have more risk to be infected and prevalence of COVID-19 in male of Japan is higher than female. While COVID-19 spread more by cases infected with in the country through community transmission as compared to foreign visitors. Second asymptomatic patients rate is almost 10%, which is hard to diagnose them only on clinical bases so laboratory based screening of people at large scale has recommended to contain and reduce the currents sudden rise of cases. ^20^

## Data Availability

All data are available at Ministry of Health, Labour and Welfare. https://www.mhlw.go.jp/index.html.

## Authors Contribution

Muhammad Qasim: Conceptualization, Methodology, Formal analysis, Investigation Supervision, Writing - original draft. Muhammad Yasir: Formal analysis, Investigation. Waqas Ahmad: Methodology, Investigation. Minami Yoshida: Data collection. Muhammad Azhar: Methodology, Formal analysis. Muhammad Azam Ali: Supervision, Chris Wang: Data collection, Maree Gould, Writing - original draft and review.

## Ethical Approval

Study does not involve any direct experiments with human or animal

## Supplementary files

Data files of 1192 cases can be provided on request of reviewers

